# Epilepsy Socioeconomic Impact in a Tertiary Center in Brazil From the Patient Perspective

**DOI:** 10.1101/2024.07.02.24309857

**Authors:** Tayla Romão, Everton Nunes da Silva, Monica Kayo, Raíssa Mansilla, Lucas Ferraz, Isabella D’Andrea

## Abstract

**Background:** This study aimed to evaluate the direct and indirect annual costs of epilepsy from the perspective of patients with epilepsy treated at a public tertiary center situated in Rio de Janeiro, Brazil.

**Methods:** A cross-sectional cost-of-illness study was conducted, using a bottom-up approach based on interviews and records of 166 outpatients with confirmed diagnoses of epilepsy. Direct costs included expenses related to treatment, and transportation, while indirect costs encompassed productivity losses due to morbidity and mortality, assessed through the human capital approach and caregivers.

**Results:** The majority of patients in the sample had refractory epilepsy (68.1%) and were on polytherapy (43.98%). The average per capita income of the sample was USD 434,90 per month, and 28.3% of the individuals were unemployed. The total costs amounted to USD 8,243.10 per patient per year, with 76.95% attributed to indirect costs, 23.05% to direct medical costs, and 2.31% to non-medical costs. The primary cost contributors included unemployment (30.42%), caregiver expenses (22.41%), and antiseizure medications (20.30%). The majority of patients reported purchasing all their medications (62.43%). The total out-of-pocket health expenses amounted to USD 2,090.10 per patient per year, with medications accounting for 90.89% of the expenses and transportation for 9.11%.

**Conclusions:** In addition to unemployment as the main cost driver, the patients incurred catastrophic spending on medications. Even though treated in a public service, out-of-pocket health expenses made up 40.04% of the average per capita income of the sample and 12.85% of the Brazilian GDP per capita in 2021. The significant patient expenditures may contribute to poor adherence to epilepsy treatment, which can exacerbate the disease and lead to increased seizure frequency. This, in turn, reduces their ability to earn income, contributing to the rise in indirect and intangible costs.

## Introduction

Epilepsy is a chronic neurological disease that affects around 50 million people worldwide, with 85% of them residing in underdeveloped countries [1], with an estimated global cost of USD 119.27 billion [2]. The average annual cost per person with epilepsy in 2019 ranged from $204 in low-income countries to $ 11,432 in high-income countries. However, the real cost of epilepsy remains unknown in most countries, particularly in low and middle-income countries [2]. A systematic review by Allers et al. (2015) identified 22 studies conducted across 16 countries, with only 7 from low and middle-income countries [3].

Brazil is an upper-middle-income country [4] with a population of over 200 million people, with about 3 million suffering from epilepsy [5–8]. Since 1988, Brazil has implemented a universal health system known as the Unified Health System (SUS – Sistema Único de Saúde). This system is based on the principles of health as a citizen’s right and the state’s duty [9]. The SUS is the main healthcare provider for approximately 75% of the Brazilian population, while the remaining have private health insurance [9]. The treatment of epilepsy in the public system in Brazil follows the Clinical Protocol and Treatment Guidelines (PCDT), which are national guidelines that dictate which procedures and medications are available in the SUS [10].

Although there are treatment guidelines that should provide access to effective treatments when followed, there is little data on the costs of epilepsy in the country. Access to medication is still a challenge in the country, with less than 50% of the patients having access to medicines through the SUS [11].

So far, no study has evaluated the direct and indirect costs of the disease from the patient’s perspective in Brazil; studies were conducted on the burden of focal and generalized seizures in the United States, Europe, and Brazil. However, the costs for Brazil were not assessed [12–14].

Therefore, we conducted a cost-of-illness study to assess the annual costs of the illness from the patient’s perspective.

## Methods

### Objective and Study Design

The study aimed to assess the annual direct and indirect costs of epilepsy from the patient’s perspective. We conducted a cross-sectional cost-of-illness study, using a bottom-up approach through interviews with outpatients from Paulo Niemeyer State Brain Institute (PNSBI), a public tertiary center in Rio de Janeiro, RJ, Brazil. This center is a reference for refractory epilepsies and neurosurgeries.

Ethical approval for this study was obtained from the PNSBI research ethics committee by the number 39119220.1.0000.8110. Both patients and caregivers signed an informed consent form after receiving explanations about the study. We collected data directly from adult patients and their families through interviews and a customized questionnaire from January 20 to December 10, 2021. The questionnaire covered sociodemographic and clinical information, treatment-related resource usage, patient expenses, and issues related to the stigma of epilepsy. To prevent memory biases, we also reviewed one year of medical records retrospectively from July 20, 2021 until March 20, 2022. The medical records were evaluated by registration number to avoid identification of participants.

### Cost estimates

Direct costs paid by patients as medical and non-medical costs were included. Costs for the year 2021 were projected in the national currency (BRL) and then converted into the dollar currency (USD) using the Purchasing Power Parity (PPP) index. This index, established by the World Bank, allows for cross-country comparisons by factoring in the Gross Domestic Product (GDP) [15]. The Brazilian PPP conversion factor for GDP in 2021 was 2.55 [16].

### Direct Costs

The direct medical costs in this study included medications, as the procedures were covered by the institution, which is part of the SUS. The evaluation of medication costs involved referring to price tables established by the Chamber of Regulation of Medicines Market (CMED). Additionally, a value equivalent to the Brazilian Tax on Circulation of Goods and Services at a rate of 20% applicable to the State of RJ was incorporated into the analysis [17].

Direct non-medical costs were associated with transportation. For public transportation, the unit cost for each ticket was considered, while for private transportation, the distance traveled in kilometers (km) was multiplied by the average price per km based on the values of services of private transportation apps. Moreover, the cost of companions was also included.

### Indirect costs

Indirect costs pertain to the loss of production due to morbidity and mortality. The Human Capital Method (HCM) is used to measure these indirect costs, which is based on the potential expected future earnings of a person if they were available to work [18].

The average gross monthly income for active individuals in the market in RJ between 2020 and 2021 was used in this study [19]. Indirect costs in this study were divided into labor market groups, namely the employed, unemployed, and incapacitated individuals.

For employed individuals, absenteeism and presenteeism were taken into account. For the unemployed, the study assessed the extent to which epilepsy contributed to their inability to secure employment on a scale from 0 (no contribution) to 10 (complete contribution). For participants unable to work, the study considered the productivity loss arising from disability retirement or the receipt of welfare sick pay due to epilepsy.

Additionally, costs associated with caregivers were included, and the evaluation encompassed the remuneration that such individuals would typically receive if they were engaged in employment in the State of RJ in 2021 [20].

## Results

### Sample characteristics

Between October 2020 and October 2021, a total of 166 patients were interviewed at the epilepsy center of PNSBI. Table 1 displays the sociodemographic profile of the study sample, showing a mean age of 36.8 (SD ± 12.7) years. The majority of participants (69.9%) relied solely on the public healthcare system for medical services. The mean per capita income was observed to be USD 434.90 (SD ± 355.69), and only 4 patients indicated having discernible income sources. In terms of their occupational status, 49.4% of the sample (n=82) were categorized as being outside the workforce (e.g. housewives, students, retirees, and people who are not interested or able to work), 28.3% (n=47) as unemployed, and 22.3% (n=37) as employed. Among the 82 individuals who were not actively part of the workforce, over half (n=54) form of financial assistance, with only 16 beneficiaries qualifying for social security retirement benefits due to their epilepsy-related condition.

**Table 1.** Sociodemographic characteristics.

| | N | Frequency (%) | Mean ±SD | Minimum-Maximum |
| --- | --- | --- | --- | --- |
| **Age (year)** | 166 | | 36.8±12.7 | 18-83 |
| **Gender** |  |  |  |  |
| Female |  | 77 (46.4%) |  |  |
| Male |  | 89 (53.6%) |  |  |
| **Education** | 166 |  |  |  |
| Illiterate |  | 16 (9.6 %) |  |  |
| Middle school |  | 42 (25.3 %) |  |  |
| High school |  | 85 (51.2 %) |  |  |
| University degree |  | 23 (13.9 %) |  |  |
| **Number of people in the household** | 166 | | 3.05±1.35 | 1-10 |
| **Family income**<br>USD/month* | 162 | | 1121.96±632.94 | 392.16-3450.98 |
| **Per capita income**<br>USD/month * | 162 | | 434.90±355.69 | 65.49-2352.94 |
| **Private health insurance** | 166 |  |  |  |
| yes |  | 50 (30.1 %) |  |  |
| not |  | 116 (69.9 %) |  |  |
| **Occupational status** | 166 |  |  |  |
| Employed |  | 37 (22.3%) |  |  |
| Unemployed |  | 47 (28.3%) |  |  |
| Outside the workforce |  | 82 (49.4%) |  |  |
| Due to epilepsy |  | 16/82 (19.51%) |  |  |

\*based on Brazilian purchasing power parity (PPP) of 2.55 (2021)

The average age of disease onset was 16.4 years (SD ± 14.9), with an average disease duration of 20.5 years (SD ± 13.1). Focal epilepsy was the most common type in the sample (146/166 cases, 87.95%), with structural causes accounting for over half of the cases (114/166, 68.7%). Among the cases with structural causes, brain tumors (41/114, 36%) and mesial temporal sclerosis (25/114, 21.9%) were the most prevalent. Regarding treatment response, 68.1% of patients (113/166) had refractory epilepsy.

In terms of epilepsy surgery, 23 patients (13.86%) had undergone such interventions previously, with only 3 surgeries taking place during the study period. In terms of mental health, a significant portion of the group (95/166, 57.2%) had psychiatric disorders, with depression and intellectual disability being the most common (see Table 2).

**Table 2.** Clinical characteristics.

|  | **N** | **Frequency (%)** | **Mean±SD** | **Minimum-Maximum** |
| --- | --- | --- | --- | --- |
| **Age at first seizure (years)** | 165 | Median = 13 | 16.3±14.9 | 3-month – 62-year-old |
| **Time of illness (years)** | 165 | Median = 20 | 20.5±13.1 | 1 - 62 |
| **Epilepsy classification** | 166 |  |  |  |
| Focal |  | **146** |  |  |
| Generalized |  | 19 |  |  |
| Unknown |  | 1 |  |  |
| **Etiology** | 166 |  |  |  |
| Structural |  | **114 (68.7 %)** |  |  |
| Unknown |  | 31 (18.7 %) |  |  |
| Genetic |  | 11 (6.6 %) |  |  |
| Infectious |  | 8 (4.8%) |  |  |
| Immune |  | 2 (1.2%) |  |  |
| **Cause of structural etiology (n=114)** |  |  |  |  |
| Brain tumor |  | **41 (36.0%)** |  |  |
| Epilepsy associated tumors |  | 16/41 (39.0%) |  |  |
| Mesial Temporo Sclerosis |  | 25 (21.9%) |  |  |
| Malformations of cortical development |  | 19 (16.7%) |  |  |
| Cerebrovascular disease |  | 15 (13.2%) |  |  |
| Hypoxic ischemic encephalopathy |  | 9 (7.9%) |  |  |
| Traumatic brain injury |  | 3 (2.6%) |  |  |
| Arachnoid cyst |  | 2 (1.8%) |  |  |
| **Psychiatric comorbidities** | 166 | 95 (57.2%) |  |  |
| Intellectual disability |  | **32/95 (33.7 %)** |  |  |
| Depression |  | 28/95 (29.5 %) |  |  |
| Anxiety disorders |  | 20/95 (21.1 %) |  |  |
| Psychogenic Nonepileptic seizures |  | 13/95 (13.7 %) |  |  |
| Dementia |  | 1/95 (1.1 %) |  |  |
| Schizophrenia |  | 1/95 (1.1 %) |  |  |
| **Refractory epilepsy** | 166 |  |  |  |
| no |  | 53 (31.9 %) |  |  |
| yes |  | **113 (68.1 %)** |  |  |

### Direct Medical Costs

In terms of the number of antiseizure medications (ASM), 16.87% (n=28) of the patients used only one, 38.55% (n=64) used two, and 43.98% (n=73) used 3 or more. The most commonly used ASMs were Clobazam (n=96), Lamotrigine (n=59), and Valproic Acid (n=40). The majority of patients (62.43%) reported that they purchased all their medicines (n=103), while only 7.27% (n=13) acquired all of them through a public source, and 30.30% (n=50) used both sources. Figure 1 illustrates the relationship between the number of patients using each ASM and the supplying source of the sample.

**Figure 1.**
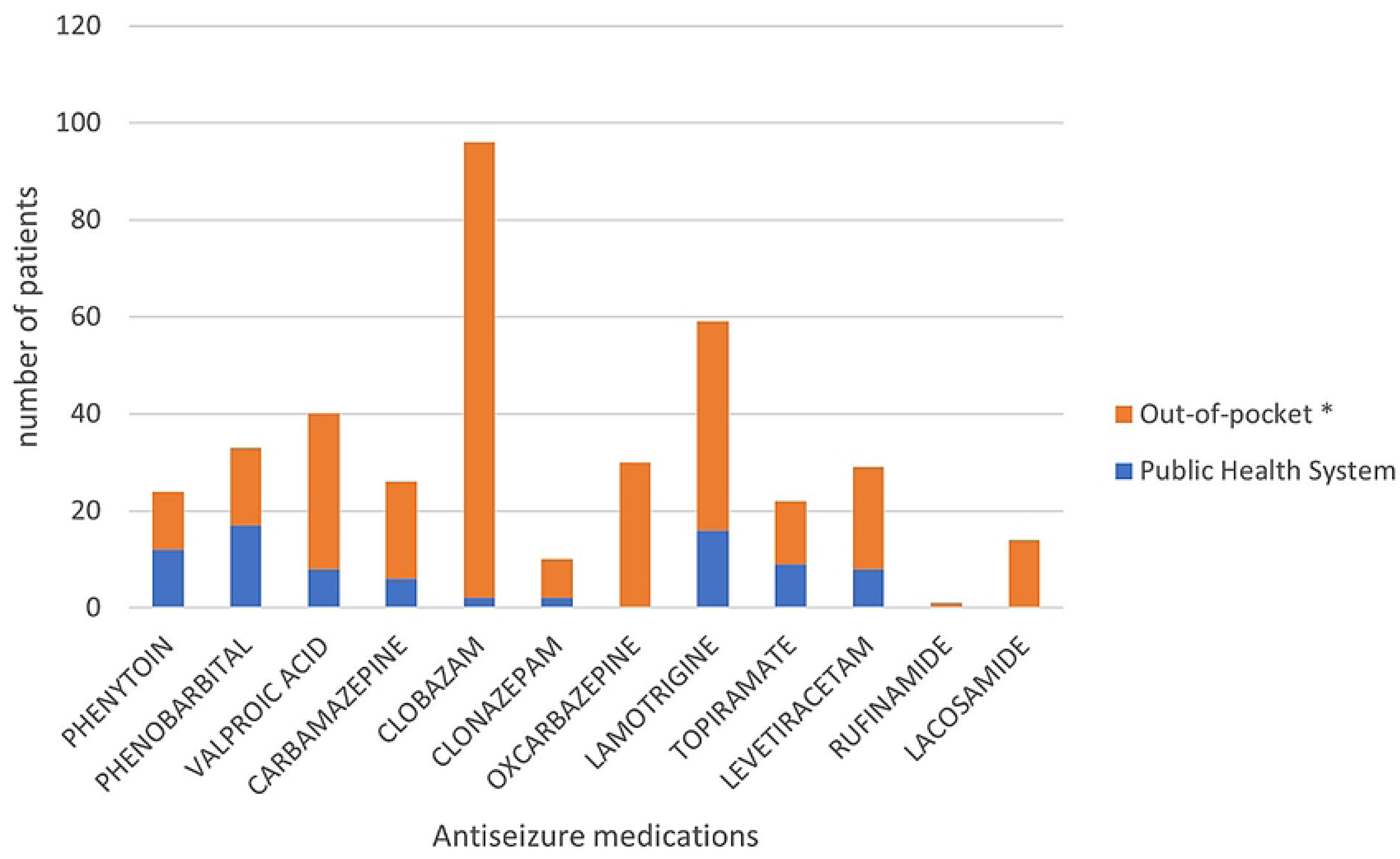
Comparison of the number of patients using various antiseizure medications and their supplying source *SUS: Unified Health System; ** refers to individuals’ direct expenses to health.

Figure 2 shows the relationship between the costs of each ASM and the supplying source. Lamotrigine is the ASM responsible for the highest costs. The main providers of ASMs were the patients, except for Phenobarbital. The total cost of ASMs was USD 330,322.14/year, with USD 277,779.31/year (84%) paid by the patients themselves.

**Figure 2.**
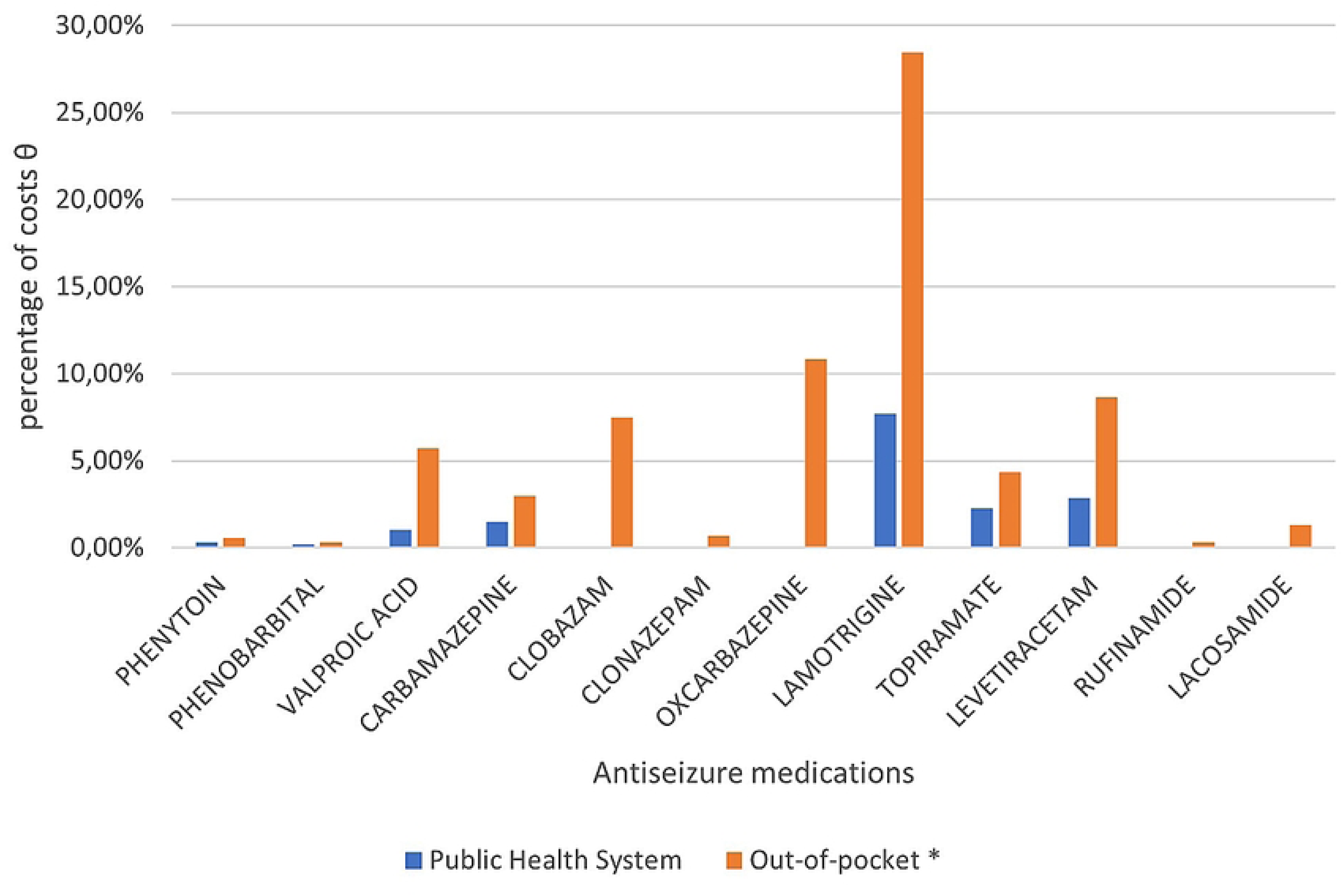
Comparison between the antiseizure medications costs and the supplying source. N=166. Total cost: USD 330,322.14/year, costs paid by patient: USD 277,779.31/year.

Regarding the costs of medications for psychiatric conditions (N=56), including antidepressants and antipsychotics, the total annual cost of mental health medications was USD 38,970.55. Of this amount, USD 37,583.55 (96.44%) was out-of-pocket.

The total annual cost of all medications, including those for mental health, was USD 369,292,69. Patients covered USD 315,362.86 (85.39%) of this cost (see Table 3). The mean out-of-pocket expense for medications was USD 1,899.78 per patient per year.

**Table 3.** Annual direct and indirect costs from the epilepsy center of Paulo Niemeyer State Brain Institute, Rio de Janeiro, Brazil.

| TYPE OF COST | N | Monetary value<br>USD* | % total<br>(indirect and<br>direct) cost |
| --- | --- | --- | --- |
| **Direct Medical Costs (out-of-pocket)** |  |  |  |
| Antiseizure medication (ASM) | 166 | USD 277,779.31 | 20.30% |
| Medications for mental disorders | 56 | USD 37,583.55 | 2.75% |
| Total Direct Medical Costs |  | USD 315,362.86 | 23.05% |
| **Direct Non-medical Costs (out-of-pocket)** |  |  |  |
| Transportation | 121 | USD 31,594.00 |  |
| Total Direct Non-Medical costs |  | USD 31,594.00 | 2.31% |
| **Indirect costs** |  |  |  |
| Caregiver | 39 | USD 306,677.65 | 22.41% |
| Absenteeism | 5 | USD 30,304.80 | 2.21% |
| Presenteeism | 21 | USD 88,941.18 | 6.50% |
| Early retirement and sick pay due to epilepsy | 16 | USD 210,823.53 | 15.41% |
| Unemployment due to epilepsy | 47 | USD 416,244.71 | 30.42% |
| Total indirect costs |  | USD 1,052,991.86 | 76.95% |
| **Total (indirect and direct) costs** | 166 | USD 1,368,354.72<br>(USD<br>8,243.10/patient/year) | 100.00% |

\* based on Brazilian PPP\* 2.55 (2021)

### Direct non-medical costs

A total of 163 patients indicated the need for transportation to attend appointments. The combined expenditure on public transportation for patients (84 individuals) and their companions (46 individuals) amounted to USD 2,123.14 per day. Furthermore, the total spent on private vehicles (utilized by 79 individuals) was USD 12,046.27 per day, with an average distance traveled of 81.95 km (ranging from 10 to 577 km).

Considering that patients had an average of 5.1 consultations per year, the total cost of transportation was USD 72,274.00 per year for the 163 patients. Of these, 121 patients reported personally covering transportation expenses, totaling USD 31,594.00 per year (as shown in Table 3). The out-of-pocket cost for transportation was USD 261,11 per patient per year.

### Indirect costs

#### Caregivers

In the sample, 39 out of 166 patients (23.49%) required caregivers, and nearly 90% of them (35 patients) had a psychiatric disorder, with intellectual deficit (21 patients) and mood disorders (9 patients) being the most common. All caregivers identified were unregistered. The estimated cost of the caregivers’ lost potential productivity was USD 306,677.65 per year (see Table 3).

#### Work absenteeism and presenteeism

Out of the 37 employed patients, 5 reported missing work due to epilepsy. The number of days they were absent ranged from 2 to 60 per year, with an average of 13.8 days, costing a total of USD 30,304.80 per year. Additionally, 40.5% (21 patients) reported reduced productivity (presenteeism) while at work due to epilepsy, with productivity decreasing by 10 to 80% (with a mean reduction of 45.3%), resulting in a total value of USD 88,941.18 per year (see Table 3).

#### Unemployment

Of the 47 unemployed individuals, 82% (n=39) stated that their inability to secure employment was due to epilepsy, with an average perceived causal relationship of 81%. The estimated cost incurred was USD 416,244.70 per year (Table 3). Some patients have reported losing job opportunities due to their epilepsy, and others have been fired from their jobs following epileptic seizures at work.

#### Early incapacity for work (early retirement)

The total cost of incapacity work due to epilepsy (n=16) was USD 210,823.53/year (Table 3).

### Total costs

The total costs from the patient’s perspective were USD 8,243.10 per patient per year (n=166). Of this amount, 76.95% were indirect costs, 23.05% were direct medical costs, and 2.31% were non-medical costs (See Table 3). When considering each cost separately, the highest expenses were for unemployment (30.42%), followed by caregiver expenses (22.41%), and ASM (20.30%).

Concerning out-of-pocket health expenses, a yearly amount of USD 2090.10 per patient (n=166) was identified, with medications accounting for 90.89% of the expenses and transportation for 9.11% (Table 4). Additionally, considering that the average per capita income of the sample was USD 434.90 per month, the out-of-pocket health expenses represented 40.04% of it.

**Table 4.** Out-of-pocket costs.

| **Out-of-pocket costs** | **N** | **USD/patient/year** | **%** |
| --- | --- | --- | --- |
| Medications | 166 | USD 1,899.78 | 90.89% |
| Transportation | 121 | USD 261.11 | 9.11% |
| **Total** |  | USD 2,090.10 | 100% |

## Discussion

Because Paulo Niemeyer State Brain Institute is a reference center for refractory epilepsy and neurosurgery, the majority of patients in our study had drug-resistant epilepsy, which comprised 113 out of 166 patients (68%). This profile aligns with the 30% global prevalence of drug-resistant epilepsy [21]. The most common type of epilepsy observed in our study was focal epilepsy of structural etiology, in contrast to the most prevalent type worldwide, which is generalized epilepsy of unknown etiology [22]. We also found that 52.7% of the patients (95 individuals) had mental disorders. This finding supports existing literature, which reports a higher prevalence of psychiatric comorbidities in epilepsy patients [23].

The unemployment rate in our sample was higher than the national average, in a period that was marked by the crisis triggered by the Covid 19 pandemic [24].

Regarding per capita income, the average gross monthly income in Brazil between 2020 and 2021 was about USD 901.96, which is twice the per capita income of our profile (USD 434.90/month). This situation is likely due to the high number of refractory patients in our profile, who often encounter difficulties with work. As a result, this could lead to underemployment or early retirement due to incapacity.

Thirty percent of our sample had private health insurance, replicating the percentage found at the national level [9,25].

The United Nation’s sustainable development goal (SDG) 3 (“Ensure healthy lives and promote well-being for all at all ages”) emphasizes the importance of achieving universal health coverage, including financial risk protection, access to quality essential healthcare services, and access to effective and affordable essential medicines [26].

However, access to medications within the SUS is still insufficient and unequal in different regions of the country, with only 45.3% of the population having full access through free distribution. Medications are often unavailable, and patients do not have the resources to purchase them, leading them to abandon their treatment, or not buy food or pay the bills [11]. Out-of-pocket spending on medicines is the main cause of impoverishment for vulnerable families, which shows us the importance of promoting access to medication as a form of financial protection, as a means of promoting SDG 1 (“End poverty in all its forms everywhere”) [26].

Our data showed that patients’ out-of-pocket spending constitutes catastrophic health spending, reaffirming the fact that in Brazil, catastrophic health spending is more common than in other countries, despite having universal health coverage [27]. The out-of-pocket financing of ASM in our sample was enormous, corresponding to 84% of ASM costs. This means that 20.3% of the total cost of treatment is financed by the patients themselves. The fact that only 22.3% of the patients were employed and the mean annual cost with ASM was USD1,899.78, even when we take into account the family income, the expenditure can still be classified as catastrophic.

The Public Health System in Brazil provides medications free of charge including the ASMs, except for Oxcarbazepine, Rufinamide, and Lacosamide. The cost of Lamotrigine accounted for the highest percentage (36.15%) of the total cost of ASMs. Despite Lamotrigine being provided for free, most patients (43 out of 59) reported having to pay for it. When questioned, many patients in our sample claimed that they have to buy their ASMs because of the frequent unavailability of them in public pharmacies and difficulty accessing them due to bureaucracy. A few declared a lack of knowledge that some of the medications were provided by the Government. A Brazilian study conducted by Boing et al. in 2013, which used a bottom-up approach (n=19,427), showed that less than half of the sample (45.3%) were able to have access to medications provided by the Brazilian Public Health System [11]. In our sample, 62.43% of patients were compelled to purchase their medications, thereby compromising their income.

Psychiatric comorbidities, which are common in epilepsy, increase the costs of the treatment. The annual cost for psychiatric medications amounted to USD 38,970.55, with 96.44% of these costs directly incurred by patients through out-of-pocket spending.

Transportation constituted 9.11% of the total out-of-pocket expenses, which could pose a financial burden on patients and families, given the high unemployment rate in our sample.

Our study was the first to analyze the indirect costs of epilepsy in Brazil. The total annual indirect costs in our sample were USD 1,052,991.86, representing 76.95% of the total costs. Unemployment was the primary cost driver, accounting for 30.42% of the costs. This aligns with a study by Libby et al, which demonstrated a productivity loss of USD 9,504.00/year for individuals with epilepsy compared to those with other chronic diseases. Their study showed that only 42% of individuals with epilepsy over 18 years of age were employed, in contrast to the 70% employment rate among people without epilepsy. They concluded that lost wage-based productivity associated with epilepsy was nearly equal to combined wage losses associated with diabetes, depression, anxiety, and asthma together [28]. Factors such as clinical challenges resulting from the disease, associated psychiatric comorbidities, adverse effects of treatment, and cognitive impairments contribute to this disparity in employment rates. Additionally, the stigma surrounding epilepsy, where 30% of individuals report experiencing discrimination due to their condition, also plays a significant role [29]. These clinical and social challenges together create substantial barriers for individuals with epilepsy to access the job market.

The caregiver was the second main cost driver, totaling $306,667.65 per year (N= 39), which corresponds to 22.41% of the total costs. All the caregivers in our study were informal, and the costs found reflected the income that they could have earned if they were working instead of providing care. Caregivers also experience negative effects on their health and psychological well-being, which represent intangible costs that were not evaluated in the current study [30].

The GDP per capita in Brazil was USD 16,260.10 (USD PPP in 2021). Out-of-pocket costs amounted to 12.85% of it (USD 2,090.10/patient/year) and indirect costs to 39% of it (USD 6,343.32/patient/year).

Our study has some limitations. The overrepresentation of patients with refractory epilepsy in the sample might limit the generalizability of the findings due to potential selection bias. It’s important to note that costs associated with neuromodulation, Cannabidiol, and epilepsy-specific diets were not quantified, which could lead to an underestimation of the overall economic burden. Additionally, the study did not consider the potential impact of the COVID-19 pandemic on costs, patient access, and economic stability.

## Conclusions

Despite the healthcare system with universal coverage, people with epilepsy have catastrophic out-of-pocket healthcare costs. It is crucial to ensure access to medications already listed in the national guidelines to protect vulnerable individuals from further impoverishment and promote equality in healthcare.

## Data Availability

All relevant data are within the manuscript and its Supporting Information files.

